# Genetically informed search for potential osteoarthritis drug targets across the proteome

**DOI:** 10.64898/2026.02.10.26345885

**Authors:** Weijie Liu, Benjamin P. Zuckerman, Art Schuermans, Gisela Orozco, Michael C. Honigberg, John Bowes, Terence W O’Neill, Sizheng Steven Zhao

## Abstract

**Background:** Osteoarthritis (OA) is a leading cause of disability worldwide, yet no licensed therapies can prevent or slow its progression. We aimed to identify potential targets for disease-modifying OA drugs (DMOADs) by integrating genetic and differential protein expression (DPE) evidence.

**Methods:** We evaluated genetically predicted perturbations of plasma protein levels using cis-protein quantitative trait loci (cis-pQTLs) across three large European cohorts (UK Biobank Pharma Proteomics Project, deCODE, and Fenland) and outcome data from the Genetics of Osteoarthritis Consortium, covering 11 OA phenotypes. DPE analyses were performed in 44,789 UKB participants, comparing 2,920 protein measurements between OA cases and controls, supported by sensitivity analyses. Proteins identified through genetic and/or DPE approaches were further assessed in downstream analyses.

**Findings:** In total, 305 proteins showed evidence of association with OA through genetically predicted perturbations, with 81 supported by colocalisation across datasets. DPE analyses identified 605 proteins associated with at least one OA phenotype, of which 450 (74·4%) remained robust after sensitivity testing. Several novel targets were identified, including PPP1R9B, PCSK7, and ITIH4. Integration of both approaches prioritised 5 proteins, 4 of which demonstrated druggable potential, including 3 high-confidence candidates DLK1, TNFRSF9, and OGN. Downstream analyses highlighted key biological pathways and candidate compounds with potential for repurposing.

**Interpretation:** This large-scale study combines genetic and DPE evidence to prioritise candidate DMOAD targets. Findings reinforce established biology while revealing novel proteins and pathways, providing a foundation for therapeutic development in OA.

**Funding:** WL is supported by the Guangzhou Elite Project (project no. JY202314). SSZ is supported by The University of Manchester Dean’s Prize, Arthritis UK Career Development Fellowship (grant no. 23258). This work is supported by the NIHR Manchester Biomedical Research Centre (NIHR203308).

**Research in context:** *Evidence before this study:* Circulating proteins have been linked to osteoarthritis (OA) in observational studies, supporting their potential as biomarkers and drug targets. However, differential protein expression analyses are vulnerable to confounding and reverse causation. Mendelian randomisation (MR) studies using proteomic GWAS instruments have suggested causal roles for several circulating proteins in OA-related traits and highlighted druggable candidates. However, many analyses relied on earlier OA GWAS data (e.g., Genetics of Osteoarthritis Consortium 1·0) and smaller proteomic GWAS datasets, and typically did not integrate MR findings with large-scale differential protein expression. As a result, it remains unclear how well genetically predicted protein effects align with observed protein expression in OA, and how robust prioritised targets are when replicated across proteomic data from multiple cohorts.

*Added value of this study:* This study integrates large-scale proteomic MR and differential protein expression (DPE) analyses across multiple OA phenotypes using the largest datasets to date. By combining genetic evidence with observed protein dysregulation in population-based cohorts, we strengthen causal inference and improve robustness of target prioritisation. This approach allows us to distinguish proteins that are likely to play a causal role in OA from those that reflect downstream disease processes, and to highlight targets with greater translational relevance than identified by either method alone.

*Implications of all the available evidence:* Taken together, our findings support a causal role for a subset of circulating proteins in OA and demonstrates the value of integrating genetic and observational proteomic data for target prioritisation. Proteins supported by both MR and DPE are more likely to represent biologically relevant drivers of disease and actionable therapeutic targets. This integrated framework reduces false positives arising from confounding or reverse causation and provides a more reliable basis for drug development, biomarker discovery, and patient stratification in OA.

## Introduction

Osteoarthritis (OA) is a major cause of disability in middle-age and older adults[1]. The most frequently affected joints are the hip, knee, hand, foot and spine, although OA can affect any joint. It is estimated that symptomatic OA affects one in eight people in the US[2]. Current management strategies have focused on reducing symptoms. To date there are no licensed pharmacologic therapies that can prevent the onset or progression of OA.

Drug development in OA can be challenging, and clinical trial programmes have been largely unsuccessful to date. Most drug targets are proteins; therefore, one common approach to drug discovery is to perform hypothesis-free comparisons of circulating protein level between OA cases and controls[3,4]. High-throughput technologies that measure thousands of proteins are increasingly available. Such case-control protein association studies can help provide context for candidate targets, improve understanding of biological pathways, and prioritise target selection. However, observational associations do not always reflect causation, and the high correlation among proteins can obscure true biological signals. Moreover, large-scale proteome-wide profiling in OA remains limited by the relatively small number of cases.

Another increasingly popular approach is to use human genetic data to improve causal inference[5]. Studies have repeated shown that drugs supported by human genetic data are more than twice as likely to succeed[6]. Natural genetic variation in genes encoding protein drug targets can offer insights into the potential benefits—or risks—of targeting those proteins pharmacologically. Such Mendelian randomisation (MR) studies can add strength to observational analyses by explicitly seeking to estimate causation. Recent studies have identified numerous genetic variants associated with circulating protein levels (protein quantitative trait loci, pQTLs) using data from thousands of individuals[7–9]. Such variants can be used to evaluate the specific effects of genetically predicted levels of certain proteins on disease incidence[7,10]. MR has been used for drug discovery in OA[11] but mostly without replication across populations or proteomic technologies[12]. MR studies, when presented alone, can also be problematic because, first, its primary inference pertains to OA prevention rather than modifying existing OA; second, effect sizes can be difficult to interpret as they reflect life-long rather than shorter-duration pharmacological interventions.

We address respective limitations of each drug discovery approach by combining case-control protein association and MR designs, in the largest sample size of either approach to date, to identify and prioritise potential OA therapeutic targets.

### Patients and methods

As an overview, this study utilized data from the UK Biobank (UKB), deCODE, and the Fenland study. UKB is a population-based cohort consisting of approximately 500,000 volunteers aged 40-69 years[13] recruited from 22 assessment centres across the UK from 2006 to 2010[14]. Detailed description of all the data sources and analysis involved in this study can be found in the supplementary methods.

### Ethics

Use of UKB data was approved by the North West Multi-centre Research Ethics Committee. Analyses were conducted under application no. 72723. Ethical approval was not required for analysis using summary-level data.

#### Drug target cis-MR and reverse MR

We used genetic association data for circulating protein levels from three independent sources: 1) UK Biobank Pharma Proteomics Project (UKB-PPP)[9]: 2,941 assays covering 2,922 unique proteins measured with the Olink Explore 3072 platform in 54,219 participants (mean age 56·2 years, SD 8·2; 53·9% female). 2) deCODE (Iceland)[8]: 4,907 assays of 4,672 unique proteins using the SomaScan v4 platform (SomaLogic, Inc.) in 35,559 participants (mean age 55 years, SD 17; 57% female among those with plasma samples). 3) Fenland (UK)[7]: 4,923 assays of 4,665 unique proteins using SomaScan v4 in 10,708 participants (mean age 48·6 years, SD 7·5; 53·3% female). Only European ancestry data from these 3 data sources were used in the analysis.

We selected single-nucleotide variants (SNVs) as instrumental variables to proxy protein perturbation in each dataset. Specifically, we selected significant (p < 5 × 10^−6^) and minimally correlated (r² < 0·1, 10 kb window, using the 1,000 Genomes phase 3 v5a reference panel[15]) SNVs from the cis-region (± 200 kb of the encoding gene), with minor allele frequency (MAF) > 0·1%. SNVs within the Major Histocompatibility Complex (MHC) region (chr 6: 26–34 Mb) were excluded. We prioritised cis-pQTLs because trans-pQTLs are more likely to be pleiotropic. To reduce bias from reverse causation (i.e., false positive associations due to OA affecting protein level), we performed Steiger filtering[16], that is, excluded pQTLs that explained greater variance in OA than protein level. F-statistics were calculated to assess potential weak instrument bias.

Outcome data were selected from the largest genome-wide association meta-analysis (GWA-meta) of OA to date (Genetics of Osteoarthritis Consortium 2·0 [GO 2·0]), comprising approximately 1·9 million individuals (489,975 cases), across 11 OA phenotypes, which includes All OA, Knee and Hip OA, Hip OA, Knee OA, Total joint replacement (TJR), Total hip replacement (THR), Total knee replacement (TKR), Spine OA, Hand OA, Finger OA and Thumb OA[17]. OA was defined using a combination of self-report, clinical diagnosis, International Classification of Diseases (ICD) codes, radiographic evidence and/or joint replacement, in each study within the meta-analysis as detailed in the original publication[17]. Controls were either OA-free or population-based, with some cohorts applying ICD code or self-report exclusions.

MR analyses were conducted for each protein from each of three data sources versus each OA phenotype. Protein levels were analysed on their original scales provided by each study without additional transformation or normalization. Therefore, genetic association estimates for the exposures represent the per-allele change in protein abundance in the native measurement units of each platform. The outcome (OA) was analysed as a binary trait (case vs control) in the corresponding GWAS datasets, and causal estimates are interpreted as the change in OA risk per genetically predicted increase in plasma protein level. In the MR analysis, we assumed that the cis-pQTL instruments were robustly associated with circulating protein levels (relevance), were not confounded by other factors (independence), and affected OA risk only through their impact on the corresponding protein (exclusion restriction). We used the Wald ratio method for single-SNV proteins, and fixed-effect inverse-variance weighted (IVW) method for multi-SNV instruments incorporating linkage disequilibrium (LD) structure to account for weak correlation. P values were adjusted for multiple testing using the Benjamini–Hochberg false discovery rate (BH-FDR) method, with correction applied independently for each data source and OA phenotype. To examine for potential genetic confounding from LD, we applied Bayesian colocalisation analysis[18] to proteins that survived multiple testing correction; that is, we assessed the posterior probability of the protein-OA association being explained by the same (rather than distinct) causal variant [18,19]. Default prior probabilities were used[19] and posterior probability of >80% was considered evidence of colocalisation.

Finally, we tested for reverse causation using reverse MR, evaluating whether the observed associations could reflect OA influencing protein levels. Independent (r^2^ < 0·001), genome-wide significant (p < 5 × 10^−8^) SNVs that passed Steiger filtering were used as instruments for each OA phenotype[16].

This study followed the Strengthening the Reporting of Observational Studies in Epidemiology using Mendelian Randomisation (STROBE-MR). ‘TwoSampleMR’ (version 0·6·9)[16], ‘MendelianRandomization’ (version 0·10·0)[20] and ‘coloc’ (version 5·2·3) R packages were used for data analysis[18,19].

#### DPE analysis

Only UKB data were used in this analysis. Olink quantifies proteins on a log-2 scale relative to controls (i.e., normalized protein expression (NPX)), thus a 1-unit increase represents a doubling of the protein level[9]. Proteins with >30% missing data were excluded, as a high proportion of missing results may indicate poor assay-quality for that specific protein.

We identified OA cases and controls among participants for whom Olink proteomic data were available. OA was defined as prevalent cases, that is, individuals with OA through self-report, ICD code[21], and/or primary care Read code, and/or OPCS code[22] before the date of the blood sample. Remaining participants were classified as the control group. Individuals with >30% missing protein levels were excluded, as this potentially indicates issues with sample quality.

We estimated the association between OA status and plasma protein levels using linear models, with protein level as the dependent variable, and OA case-control status as the main explanatory variable. The model was adjusted for covariates including age, sex, and body mass index (BMI, kg/m^2^). Sensitivity analyses included additional covariates beyond the primary analysis: the top three genetic principal components (PCs), smoking status (ever/never), and the Townsend Deprivation Index (TDI). P-values were corrected to account for multiple testing within each OA phenotypes.

Missing protein levels were imputed using the K-nearest neighbour method (k = 10). For BMI, values originally derived from measured height and weight were first replaced with impedance-based estimates where available. Remaining missing values for BMI, smoking status (ever/never), and TDI were then singly imputed using chained equations. The imputation model included age, sex, and the top 10 genetic principal components as auxiliary variables.

Given the large sample size, we anticipated that even associations of negligible magnitude could reach statistical significance. To address this, we estimated a minimum detectable difference (MDD) based on the lowest intra-sample assay coefficient of variation (CV) and excluded regression coefficients < 0·051. BH-FDR correction was applied separately per OA phenotype.

#### Biological insights

To further prioritise and provide functional context for candidate proteins, we performed a series of analyses using proteins identified from MR and differential protein expression (DPE) analysis respectively.

##### Over-representation analysis

To gain insights into their biological functions, we performed Gene Ontology (GO) over-representation analysis [23], which categorizes potential function relevance to Molecular Function (MF), Cellular Component (CC), and Biological Process (BP) [23]. We also contextualized the candidate proteins within known signalling and metabolic pathways using WikiPathways 2024[24]. We then performed drug over-representative analysis using the Drug Signature Database (DsigDB) by querying the set of candidate proteins to identify approved drugs and small molecules with known or predicted interactions[25].

##### PPI network

We sought proteins most central to shared pathways by examining protein-protein interaction (PPI), which provides comprehensive information on both physical and functional protein associations, to explore the potential biological interconnectivity among these proteins. The network was built using STRING (Search Tool for the Retrieval of Interacting Genes)[26], and centrality measures were calculated with the CytoNCA app[27] in Cytoscape (v3·10·3).

##### Druggability assessment

We assessed also how likely that candidate proteins can be targeted therapeutically, using a previously published list of druggable genes[28]. Genes were classified into 3 tiers based on the strength of evidence that a gene encodes a protein target with druggability, reflecting its current or potential use in therapeutic intervention. Tier 1: approved drug targets and targets of drugs in clinical development; Tier 2: proteins closely related to Tier 1 drug targets or with associated drug-like compounds; Tier 3: incorporated extracellular proteins and members of key drug-target families.

### Role of funders

The funding source had no role in the study design; in the collection, analysis, or interpretation of data; in the writing of the manuscript; or in the decision to submit the manuscript for publication. All co-authors were not precluded from accessing data in the study, and they accept responsibility to submit for publication.

## Results

Primary results from this study are summarised in Supplementary Table S1.

### MR analysis

As outlined above, the exposure datasets included 54,219 (UKB-PPP), 35,559 (deCODE), and 10,708 (Fenland) participants of European ancestry, and the outcome data were derived from the GO 2·0 meta-analysis comprising approximately 1·9 million individuals (489,975 OA cases) across 11 phenotypes. Heterogeneity metrics for the contributing cohorts were not reported in the original OA meta-analysis. The exposure datasets included only European ancestry participants, while the outcome GWAS comprised predominantly European individuals (≈ 87·3%), ensuring broadly comparable genetic backgrounds. Participant overlap was minimal (< 0·1% for UKB-PPP and deCODE, Fenland).

From the cis-MR analyses, we identified 81 unique proteins causally associated (colocalized) with at least one OA phenotype (Supplementary Table S2): 34 from UKB-PPP, 39 from deCODE, and 47 from Fenland (detailed list in Supplementary Table S3). Across all OA phenotypes combined, 28 proteins-OA associations were replicated in at least one other data source with 11 (including PCSK7) replicated in all three. Nine from UKB-PPP and 2 from each of deCODE and Fenland could not be replicated due to their unique presence in each data source (Supplementary Fig. S1; Supplementary Table S4). Forty proteins replicated in at least one other OA phenotype, while no proteins replicated in all 11 OA phenotypes (Supplementary Table S5 and Supplementary Fig. S2). Fig. 1 shows Miami plots of protein associations with all-OA in each of the protein data sources; equivalent plots for specific OA traits are shown in supplementary Fig. S3-5.

**Figure 1.**
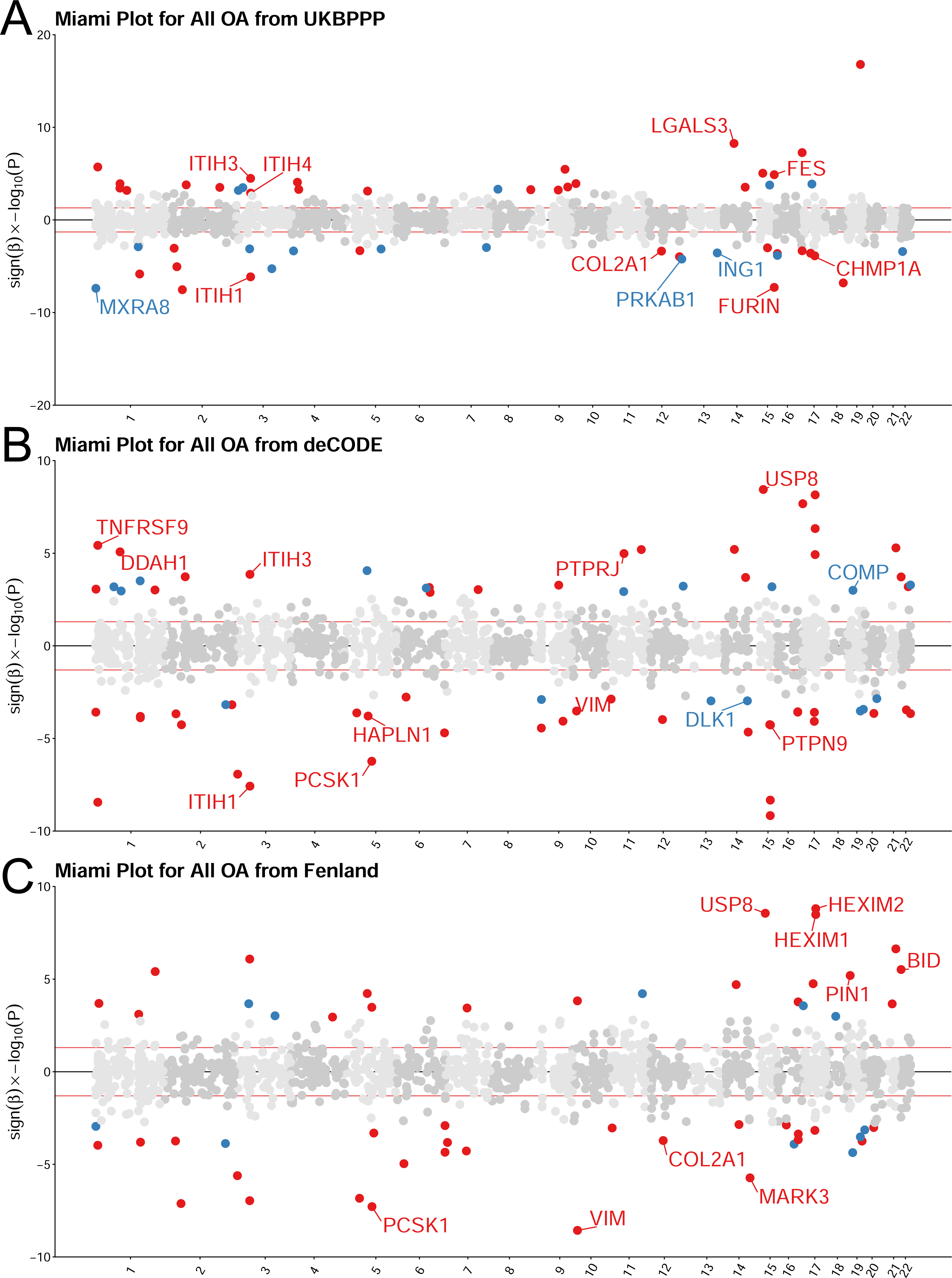
Miami plot showing the results of all OA trait from MR analyses based on UKB-PPP, deCODE and Fenland (Fig. 1A-1C). The x-axis represents the chromosomal position of each protein’s encoding gene, while the y-axis shows the signed beta multiplied by the negative log_10_-transformed p-value (after B-H procedure), allowing visualization of effect directions. The two red horizontal lines mark the significance threshold (p = 0·05). Proteins in blue show MR evidence only in the current OA phenotype, whereas those in red are supported by MR evidence in two or more OA phenotypes. Proteins that meet colocalisation criteria (PPH_4_ > 0·8) are labelled with their protein names. OA, osteoarthritis; MR, mendelian randomisation; UKB-PPP, UK biobank Pharma Proteomics Project; B-H, Benjamini-Hochberg; PPH_4_, posterior probability of hypothesis 4.

Among the candidate proteins, Protein polybromo-1 (PBRM1) showed the strongest association (by p-value) with knee and hip OA (Wald ratio: OR = 1·6, 95% confidence interval [CI]:1·4-1·8; *P* < 4 × 10^−16^), followed by Matrix Gla protein (MGP) with finger OA (OR = 0·8, 95% CI: 0·7-0·8; *P* < 1 × 10^−15^) and Inter-alpha-trypsin inhibitor heavy chain H3 (ITIH3) with TJR (OR = 1·2, 95% CI: 1.1-1.2; *P* < 1 × 10^−14^; Supplementary Table S1). Notably, both MGP and ITIH3 replicated in at least one other data source or OA phenotype.

All instruments had F-statistics > 20 (Supplementary Table S6). No instrument variables (IVs) were removed in Steiger filtering in reverse MR (Supplementary Table S7) and we found no evidence of reverse causation (Supplementary Table S8).

### DPE analysis

We identified 605 unique proteins associated with at least one OA phenotype after multiple testing correction and MDD filtering. The number of proteins varied by phenotype, from 297 for spine OA to one for the hand OA. Results for each OA phenotype are shown in Supplementary Table S9.

Among these results, Collagen alpha-1(IX) chain (COL9A1) showed the most significant signal with all OA, followed by Kallikrein-4 (KLK4) with knee and hip OA and Chromogranin-A (CHGA) with all OA. Fig. 2 summarises results for all OA in a volcano plot, with corresponding volcano plots for the other seven OA phenotypes shown in supplementary Fig. S6). Most associations (450 / 605; 74·4%) were replicated in sensitivity analysis adjusting for additional covariates (Supplementary Table S10).

**Figure 2.**
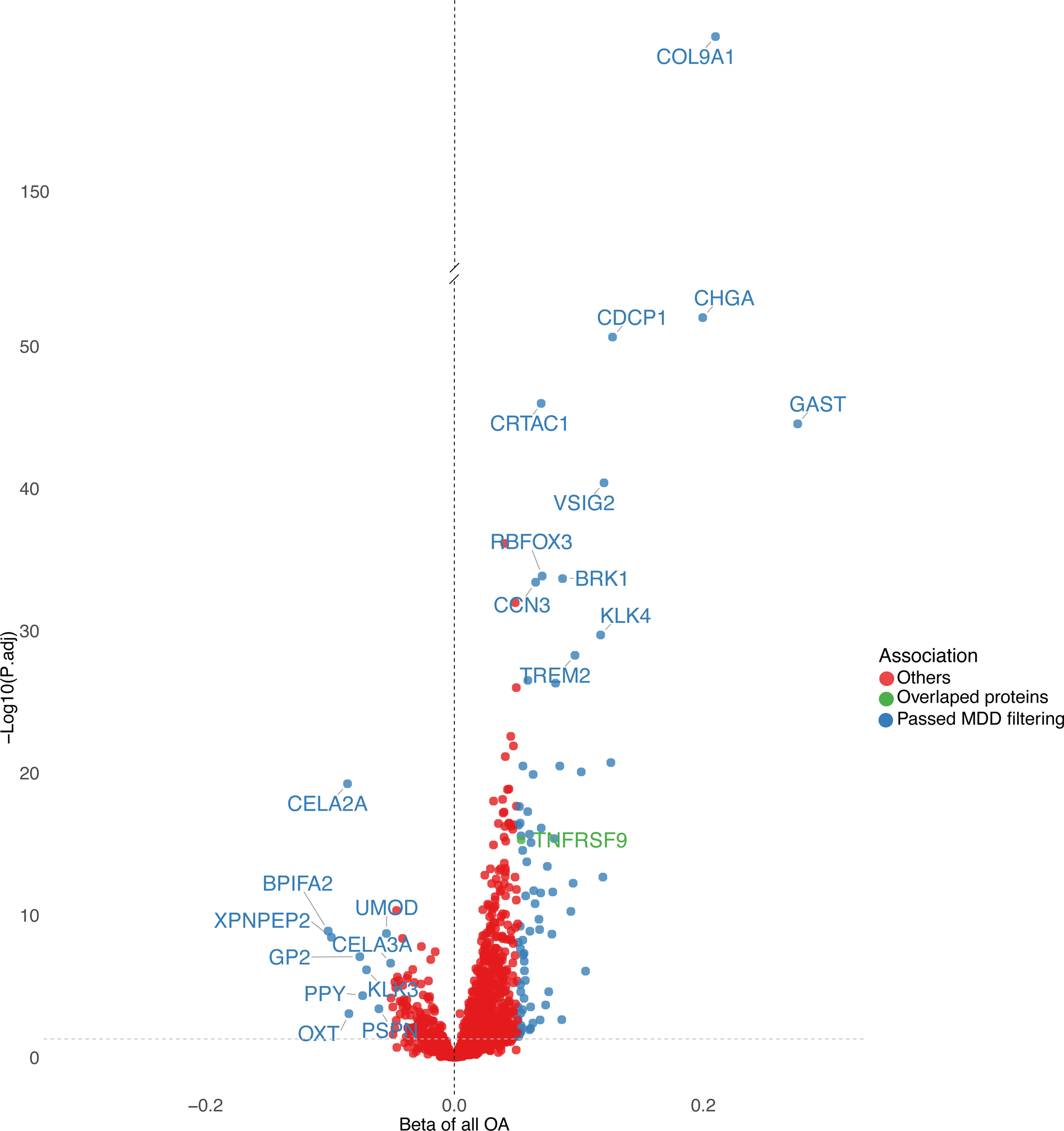
Volcano plot of the DPE analysis for overall OA. The x-axis shows the beta coefficients representing the effect size of each protein on OA risk, while the y-axis indicates the –log10 of the adjusted p-values from linear regression. Proteins that passed the MDD threshold (Beta > 0·051; see supplementary methods) are highlighted in blue. Proteins overlapping between DPE and MR analyses across three proteomic datasets with matched OA types are shown in green. Among the most significant proteins based on p-values, the top 10 on each side of the beta distribution are labelled. Other proteins are shown in red. DPE, differential protein expression; OA, osteoarthritis; OR, odds ratio; MDD, minimum detectable difference.

### Overlap between cis-MR and DPE analysis

When comparing matched OA phenotypes, five proteins demonstrated consistent associations across both the cis-MR and DPE analyses, with 2 of them exhibiting tier 1 druggability (Table 1). Protein delta homolog 1 (DLK1) was associated with reduced OA risk in MR and correspondingly observed at lower circulating levels in individuals with knee and hip OA, as well as knee OA (Table 1). Tumour necrosis factor receptor superfamily member 9 (TNFRSF9) was linked to increased OA risk in MR and higher circulating levels in individuals with all-OA, knee and hip OA, and hip OA. Neurabin-2 (PPP1R9B) showed a risk-reducing effect in MR but higher circulating levels in individuals with knee and hip OA. Mimecan (OGN) increased OA risk in MR and was also elevated in individuals with knee OA. Finally, the complement component C9 (C9) was associated with higher OA risk in MR and elevated levels in individuals with hip OA in the DPE analysis.

**Table 1.** Summary of 17 Proteins with MR Evidence and Tier 1 Druggability Support*.

| Protein | Available Data Sources | Data sources with MR Signals | OA Phenotypes with MR Signals† | Replication between DPE and MR‡ | PPI Center |
| --- | --- | --- | --- | --- | --- |
| DLK1 | All 3 data sources | All 3 data sources | ALLOA; KNEEHIP; KNEE; TKR | Yes | No |
| DPEP1 | All 3 data sources | deCODE; UKB-PPP | HIP; KNEEHIP; KNEE; TJR; TKR | No | No |
| PDGFRA | All 3 data sources | UKB-PPP; Fenland | THR | No | No |
| SOD3 | All 3 data sources | UKB-PPP; Fenland | HIP; KNEEHIP; THR; TJR | No | No |
| VIM | All 3 data sources | deCODE; Fenland | ALLOA; HIP; KNEEHIP; TJR | No | No |
| CSK | deCODE;Fenland | deCODE | TKR | NA | No |
| CXCL10 | All 3 data sources | Fenland | HAND | No | Yes |
| DDR2 | deCODE;Fenland | Fenland | TKR | NA | No |
| FGFR3 | deCODE;Fenland | Fenland | KNEEHIP; THR; TJR | NA | Yes |
| FGFR4 | All 3 data sources | UKB-PPP | KNEE; TKR | No | No |
| MAPK3 | deCODE;Fenland | deCODE | KNEE | NA | Yes |
| MAPK9 | All 3 data sources | deCODE | HAND | No | No |
| MARK3 | deCODE;Fenland | Fenland | ALLOA; KNEEHIP; KNEE; TJR; TKR | NA | No |
| PRKD2 | UKB-PPP | UKB-PPP | HIP | No | No |
| SOST | All 3 data sources | Fenland | THR; TJR | NO | Yes |
| TNFRSF9 | All 3 data sources | deCODE | ALLOA; HIP; KNEEHIP; THR; TJR | Yes | No |
| ULK3 | deCODE;Fenland | deCODE | TKR | NA | No |

\* Tier 1: targets of approved drugs or drugs in clinical development.
† Replication between DPE and MR; same OA phenotype though could be different MR data source.
‡ PPI centres were defined according to the eight centrality metrics (see supplementary methods).
MR: Mendelian randomisation; All 3 data sources: UK Biobank-Pharma Proteomics Project (UKB-PPP), deCODE, and Fenland; OA: osteoarthritis; UKB-PPP: UK Biobank Pharma Proteomics Project; TJR: total joint replacement; TKR: total knee replacement; THR: total hip replacement; KNEEHIP: knee and hip OA; NA: not applicable if protein does not exist in UKB-PPP; PPI: protein–protein interaction; DPE: differential protein expression.

Eight additional proteins showed evidence in both MR and DPE analyses, although for different OA phenotypes (Supplementary Table 2): pro-adrenomedullin (ADM), butyrophilin subfamily 2 member A1 (BTN2A1), cartilage oligomeric matrix protein (COMP), furin (FURIN), galectin-3 (LGALS3), sclerostin (SOST), thrombospondin-2 (THBS2) and C-X-C motif chemokine 10 (CXCL10).

### Biological insights

In the GO over-representation analysis of 81 unique MR significant proteins, protein autophosphorylation (enriched by *PDGFRA* etc.), collagen-containing extracellular matrix (enriched by collagen type XXVIII alpha 1 chain [*COL28A1*] etc.), and transmembrane receptor protein tyrosine kinase activity (enriched by platelet-derived growth factor receptor alpha [*PDGFRA*] etc.) were the most significantly enriched terms in BP, CC, and MF, respectively (Fig. 3; Supplementary Table S11). In the over-representation results of WikiPathways, the Osteoarthritic Chondrocyte Hypertrophy pathway shows the highest over-representation result, followed by the Focal Adhesion and Focal Adhesion PI3K Akt mTOR Signaling pathways. In the gene frequency plot (Supplementary Fig. S7), *PDGFRA*, fibroblast growth factor receptor 3 (*FGFR3*), fibroblast growth factor receptor 4 (*FGFR4*), discoidin domain-containing receptor 2 (*DDR2*), and MAP kinase-activated protein kinase 3 (*MAPK3*) show high frequency and are enriched in the Osteoarthritic Chondrocyte Hypertrophy pathway. Drug over-representation results from DSigDB 2024 database show that both NVP−TAE684 and HG-9-91-01 had the highest over-representation results, and the top 5 terms along with their enriched gene set are listed in Fig. 3.

**Figure 3.**
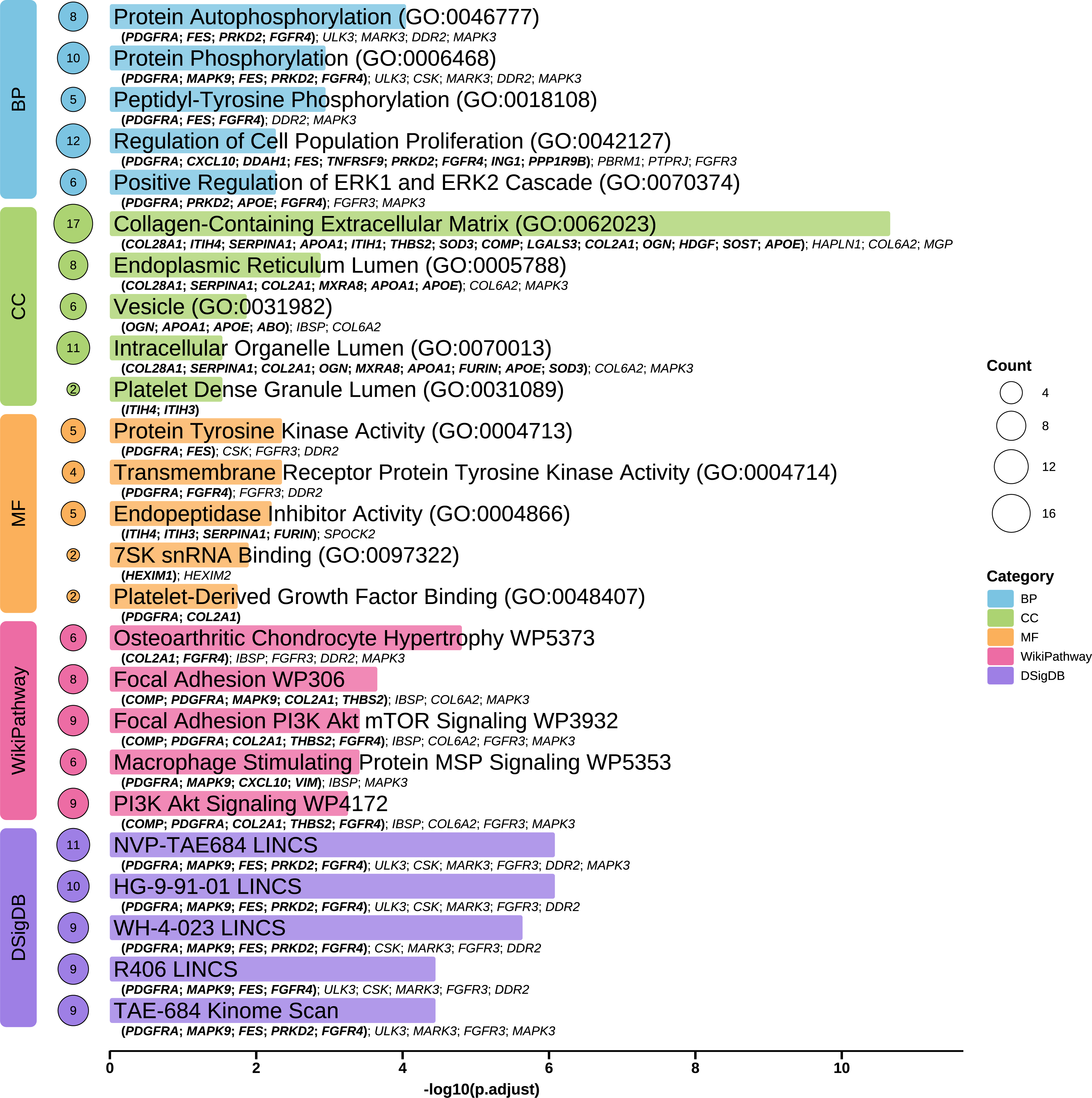
Top five over-representation analysis results based on 81 MR-significant proteins (colocalized), using GO, WikiPathway, and DsigDB. The GO consists of BP, CC, and MF. For each pathway or term, the associated proteins are listed, with those supported by both DPE and MR highlighted in bold. Circle sizes represent the number of proteins enriched in each pathway or term. GO, Gene Ontology; DsigDB, Drug Signature Database; BP, biological process; CC, cellular component; MF, molecular function; OA, osteoarthritis; DPE, differential protein expression; MR, mendelian randomisation.

The results using 605 proteins with DPE evidence and using 34 MR significant proteins from the UKB-PPP are presented in Supplementary Fig. S8 and Supplementary Fig. S9 for reference.

PPI network analysis was performed using 81 MR significant proteins (Fig. 4). The PPI results showed 79 nodes and 98 edges (expected number of edges: 51; *P* < 0·001), with proteins such as MAPK3 exhibiting the highest degree (degree = 13), indicating that it interacts with the most proteins and may function as a network hub. 14 proteins in the subnetwork were identified as centre proteins (including FGFR3, ITIH4) of the PPI after applied the median selection filtering method (Supplementary Methods). Betweenness centrality values from main PPI results suggest that MAPK3 also acts as a key connector in the network (Supplementary Table S12).

**Figure 4.**
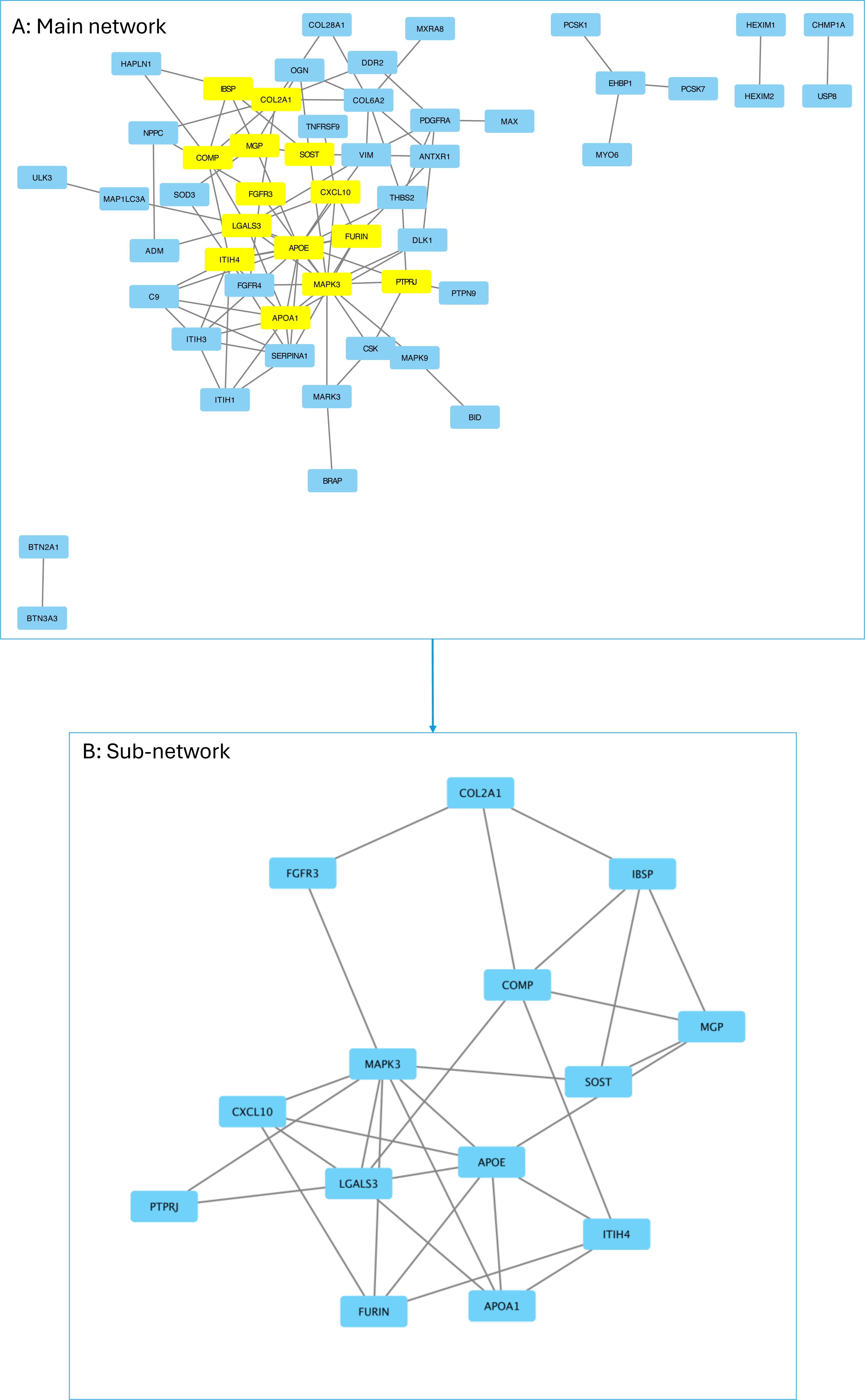
PPI network. Fig. 4A: PPI main network; proteins highlighted in yellow represent central proteins which are defined by eight centrality metrics (see supplementary methods); Fig. 4B: PPI sub-network constructed using the central proteins (as highlighted in Figure 4a). Results generated by using 81 MR significant proteins (colocalized) in the STRING database. PPI, protein-protein interaction; STRING, Search Tool for the Retrieval of Interacting Genes/Proteins.

Of the 81 proteins with MR evidence, 51 (63·0%) are considered druggable gene targets (Tier 1-3; including PCSK7 and ITIH4) (Supplementary Table S13). Among these, 17 proteins (Table 1) correspond to druggable genes that are either targets of approved drugs or targets of drugs currently in clinical development (Tier 1).

## Discussion

In the largest study of its kind, we integrated two complementary approaches commonly used in drug discovery-MR and disease-protein association analyses-to prioritise potential therapeutic targets for OA. Among these, DLK1 emerged as a leading candidate, supported by multiple lines of evidence and also highlighted in the GO 2·0 study[17]. We also identified several novel targets not previously linked to OA. Notably, many protein-OA associations mapped to biological pathways with less established relevance to OA than those detected through MR alone, underscoring the value of combining these methods in drug target research.

Among the 81 MR-significant proteins, PBRM1, MGP and ITIH3 emerged as the top candidates. Genetically predicted PBRM1 was associated with an increased risk of knee and hip OA in Fenland. PBRM1, a tumour suppressor involved in chromatin remodelling and PI3K signalling[29], is expressed in OA cartilage and lies within a risk locus, highlighting its potential as a novel DMOAD target[30]. MGP was associated with a reduced risk of finger OA and has previously been linked to reduced expression in OA cartilage and regulation of matrix mineralization[31]. ITIH3, which was associated with an increased risk of TJR in UKB-PPP and was replicated across several OA phenotypes, has been implicated in other joint diseases[32] but not OA, suggesting a potential new therapeutic direction.

Among MR-prioritised targets without prior OA evidence, PPP1R9B showed concordant prioritisation across MR and DPE, PCSK7 replicated consistently across all three proteomic sources, and ITIH4 emerged as a central PPI hub. For PPP1R9B, MR suggested that genetically predicted higher levels are associated with lower risk of knee and hip OA, while DPE showed higher circulating PPP1R9B in OA cases. Because PPP1R9B is an intracellular scaffolding protein, we interpret plasma levels as a proxy for cellular expression and/or release from damaged cells during disease. PPP1R9B anchors protein phosphatase 1 to the actin cytoskeleton and helps stabilize F-actin, a process important for maintaining cellular structure and phenotype[33–35]. This provides a plausible framework in which higher lifelong PPP1R9B activity could be protective, whereas elevated circulating PPP1R9B in established OA may represent a damage-related “leakage” signal. Future work should test whether PPP1R9B is detectable in synovial fluid and whether circulating levels track joint turnover. PCSK7 encodes a proprotein convertase that processes multiple precursors, including mediators relevant to metabolic and inflammatory signalling, and genetic and experimental studies implicate it in lipid traits and macrophage activation, supporting it as a candidate pathway for metabolic/inflammatory OA mechanisms[36]. ITIH4 is an acute-phase protein within the inter-α-trypsin inhibitor family with roles in protease regulation and extracellular matrix stabilisation; its network centrality and biology suggest a potential link between systemic inflammation, protease activity, and OA susceptibility or progression[37].

Pathway analyses revealed enrichment of Collagen-Containing Extracellular Matrix cellular function, with *COL28A1*, *COMP* (PPI centre) and *THBS2* linked to cartilage degradation and extracellular matrix (ECM) remodelling. The MAPK/ERK signalling pathway also featured prominently, with genes such as *MAPK3* (PPI centre), *FGFR3* (PPI centre), and *PDGFRA* (overlapped in association results) enriched across ERK1/ERK2 cascade[38], Peptidyl-tyrosine Phosphorylation (GO-BP)[38], PI3K-Akt-mTOR, and Focal Adhesion pathways[39]. These pathways/BP regulate chondrocyte proliferation, stress responses, and inflammation, making them promising drug targets[38,39]. In the gene frequency plot, top frequent genes like *PDGFRA*, *FGFR3* (PPI centre), and *FGFR4* are members of the receptor tyrosine kinase (RTK) family, which has been reported to promote chondrocyte hypertrophy[40]. Additionally, Protein Tyrosine Kinase Activity was identified as an enriched term under GO-MF. Collectively, our results point to tyrosine kinase–mediated activation of MAPK, PI3K–Akt–mTOR, and focal adhesion signalling cascades-converging on ECM remodelling-as a promising focus for future therapeutic development. Drug over-representation analysis highlighted NVP-TAE684, an ERK1/ERK2 inhibitor, and R406, a compound with efficacy in rheumatoid arthritis (RA)[41], as promising candidates for OA.

When restricting to matched OA phenotypes, only DLK1 and OGN showed cross-platform validation (SomaScan and Olink) with consistent effects in both association and MR analyses. In addition, of the 51 identified druggable proteins, only DLK1 and TNFRSF9 were Tier 1 targets, demonstrating consistent effects across both association and MR analyses. *DLK1* encodes a Notch/Delta family protein that regulates bone mass by inhibiting formation and promoting resorption[42]. Our findings indicate a protective effect of DLK1 across multiple OA phenotypes, with lower plasma levels observed in OA cases. Hatzikotoulas et al. also highlighted *DLK1*-expressing chondrocytes as enriched across phenotypes, suggesting cartilage as the primary tissue of action[17]. Experimental data by Berecic et al. further show enhancing *DLK1* expression induces regeneration in myocardium, raising the intriguing possibility that *DLK1* activation, rather than inhibition, could have therapeutic benefit in OA[43]. OGN, proteoglycan in particular, was reported to be upregulated in OA cartilage, as confirmed by differential gene expression and single-cell analyses [44,45]. Although our results show that OA cases have higher OGN levels compared with controls, and MR indicates that OGN increases the odds of OA, further cellular and animal studies are needed to evaluate the therapeutic potential of OGN inhibitors in OA. *TNFRSF9*, also known as CD137, encodes a transmembrane protein with a soluble form implicated in ECM metabolism[46]. We observed higher levels in OA cases, and MR suggested increased risk with higher expression, aligning with preclinical studies where TNFRSF9 inhibition was protective. Together, these results support DLK1, OGN and TNFRSF9 as high-priority therapeutic targets.

Our results aligned with prior evidence for druggable pathways, supporting face validity of the prioritisation—most notably for SOST. Romosozumab (a SOST inhibitor) has been reported to be associated with lower OA risk in osteoporosis populations[47], and in our MR analyses genetically predicted higher SOST was associated with lower risk of TKR. FGFR3 showed a similar pattern of concordance: sprifermin, which activates FGF receptors (including FGFR3), has demonstrated structural effects on articular cartilage in a randomized trial[48], and genetically predicted higher FGFR3 was associated with lower risk of knee/hip OA and joint replacement outcomes in our analyses.

The main strength of this study is the integration of multiple large proteomic datasets (UKB-PPP, deCODE, and Fenland) with UKB case–control data. There are though a number of limitations which need to be considered in interpreting the findings. First, plasma proteins may not fully represent tissue specific protein expression. This means that some biologically relevant proteins in joint tissues might not be captured, and our findings should be interpreted as reflecting systemic rather than local effects. Second, measurements were taken at a single time point, limiting insight into temporal dynamics. However, by combining cross-sectional linear regression with MR, we were still able to strengthen causal inference despite the lack of longitudinal data. Third, certain OA phenotypes (e.g. hand, finger, thumb) had relatively small sample sizes thus limiting statistical power and findings in these phenotypes should therefore be interpreted with caution, as true associations may have been missed or underestimated. Fourth, lack of sex-stratified data prevented evaluation of sex-specific effects. Fifth, DPE analyses were not possible in deCODE and Fenland due to summary-level data restrictions. This restricted our ability to directly replicate DPE associations across all cohorts, although cross-method validation between MR and UKB DPE analyses still showed consistent results. Sixth, while some sample overlap between OA and protein datasets may bias MR estimates, all instruments had F-statistics >20, reducing risk of weak instrument bias. Lastly, our data included only European ancestry participants, generalizability to other populations may be limited.

In conclusion, this study provides a broad search for candidate DMOADs by integrating proteomic and genetic data from three large cohorts. Several proteins—including DLK1, TNFRSF9, and OGN—emerge as high-priority targets, while pathway analyses highlight ECM remodelling, MAPK and PI3K signalling as central mechanisms. Novel candidates such as PPP1R9B, PCSK7, and ITIH4 warrant further wet-lab investigation. Drug enrichment results (e.g., NVP-TAE684, R406) further point to opportunities for therapeutic translation. Together, these findings expand the repertoire of promising targets for OA drug development.

## Data Availability

UK Biobank data are available to all bona fide researchers for use in health-related research that is in the public interest. The application procedure is described at www.ukbiobank.ac.uk. Summary-level protein GWAS data (UKBPPP, deCODE, Fenland) and OA GWAS data (GO 2.0 consortium) are publicly available

https://www.synapse.org/Synapse:syn51364943/wiki/622119

https://www.omicscience.org/apps/pgwas/

https://www.decode.com/summarydata/

## Contributors

All authors made substantial contributions to: (1) the conception and design of the study, acquisition of data, or analysis and interpretation of data; (2) drafting the article or revising it critically for important intellectual content; and (3) final approval of the version to be submitted. WL takes overall responsibility for the integrity of the work, from inception to the final article. SSZ has directly accessed and verified the underlying data reported in the manuscript.

## Declaration of interests

M.C.H. reports site principal investigator work for Novartis and research support from Genentech. All other authors report no relevant competing interests.

## Acknowledgement

The authors thank the participants of the UK Biobank, deCODE, and Fenland studies. The authors are grateful for the OA code list provided from the Genetics of Osteoarthritis Consortium 1·0 publication and for web resources from the Keele Health and Care Data Research Network. The authors also acknowledge the Genetics of Osteoarthritis Consortium 2·0 for providing OA GWAS data. The authors acknowledge the support of Research IT and the use of the Computational Shared Facility at The University of Manchester. SSZ is supported by an Arthritis UK Career Development Fellowship (grant no. 23258). GO, TON and SSZ acknowledge the support of Versus Arthritis (centre grants 21754 / 22084). The study has been delivered through the National Institute for Health and Care Research (NIHR) Manchester Biomedical Research Centre (BRC) (NIHR203308). The views expressed are those of the author(s) and not necessarily those of the NIHR or the Department of Health and Social Care.

## Data sharing statement

UK Biobank data are available to all bona fide researchers for use in health-related research that is in the public interest. The application procedure is described at www.ukbiobank.ac.uk. Summary-level protein GWAS data (UKBPPP, deCODE, Fenland) and OA GWAS data (GO 2·0 consortium) are publicly available.

