## supplementary methods for "Genetically informed search for potential osteoarthritis drug targets across the proteome"

**1. Data Sources and Study Design**

An overview of the study workflow is provided in Supplementary Figure S10.

***1.1 UK Biobank***

UK Biobank (UKB) is a population-based cohort consisting of approximately 500,000 volunteers aged 40-69 years[1]. Participants were recruited from 22 assessment centres across the UK, and recruitment focused on individuals living within a 25-mile (~40 km) radius of any of these centres[2]. The recruitment period spanned from 2006 to 2010[2].

We defined 11 osteoarthritis (OA) phenotypes (all OA [overall OA], knee OA, hip OA, knee and hip OA, total joint replacement [TJR], total knee replacement [TKR], total hip replacement [THR], spine OA, hand OA, finger OA and thumb OA) by using International Classification of Diseases (ICD) codes, Read codes and OPCS codes. To be specific, patients diagnosed with OA by using general practitioner (GP) records and hospital diagnoses, along with their diagnosis date and basic information (age, gender, assessment date) were collected in our study. Specifically, Read-v2 codes and CTV-3 codes from the code list published on the Keele University website were used to identify OA diagnoses from GP data[3]. The CTV-3 codes were mapped using the Read-v2 to CTV-3 code mappings from the UK Biobank's 'Coding System Lookups and Mappings Version 3' (May 2021). For hospital data, ICD-10 data from the supplementary materials of the publication published by Boer et al.[4] and the corresponding ICD-9 codes were used to find the OA diagnoses. Corresponding ICD-9 codes were mapped by using the ICD-10 to ICD-9 code mappings from the above-mentioned mapping files. OPCS codes for TJR, TKR and THR are adapted from a study by Harper et al.’s[5] (Supplementary Table S16). The OA group was defined as individuals without any OA-related diagnosis records or those who were diagnosed with OA prior to their visit to the assessment centre, where the blood sample was collected. The rest of the participants in the OLINK protein biomarkers dataset were classified as the control group.

***1.2 UK Biobank Pharma Proteomics Project***

The UK Biobank Pharma Proteomics Project (UKB-PPP) is a collaborative initiative between the UKB and pharmaceutical companies aimed at characterizing the plasma proteomic profiles of 54,219 participants. Among these participants, 46,595 UKB participants were randomly selected at the baseline visit, 6,376 individuals were selected by the UKB-PPP consortium members and 1,268 individuals were selected from the COVID-19 repeat-imaging study[6]. Those randomly selected baseline patients were highly representative for the overall UKB population across most of the demographic characteristics. Further details of the project are provided in the study by Sun et al[6].

***1.2.1 Plasma sample processing***

Plasma samples were collected and stored in EDTA vacutainers according to the pre-described UK biobank collection protocol[7]. The plasma was profiled using Olink technology, which employs Proximity Extension Assay, where a pair of antibodies labelled with unique complementary oligonucleotides (proximity probes) bind to their respective target proteins in the sample[8]. In this process, the probes come into close proximity and hybridize with each other, enabling DNA amplification of the protein signal, which is then quantified using next-generation sequencing read-out[8].

A total of 2,941 protein analytes were measured, capturing 2,923 unique proteins across 8 different panels (the Cardiometabolic I and II, Inflammation I and II, Neurology I and II, and Oncology I and II panels) using the antibody-based Olink Explore 3072 platform (Olink Proteomics, Inc; Waltham, MA)[8]. Comparisons between Olink and SomaScan measurements were made in this study and Olink assay technology (r_Olink_= 0.80-0.85) shows stronger correlations in association effect sizes for overlapping proteins compared to SomaScan (*r_SomaScan_*= 0.43-0.73).

***1.2.2 Quality control, genotyping and imputation***

Details of the quality control (QC), genotyping, and imputation procedures are provided in the study from Sun et al[6].

***1.2.3 GWAS analyses***

REGENIE v.2.2.1 was used to perform a genome wide association study (GWAS). Variants were filtered to include only those with imputation information score (INFO)> 0.7 and minor allele frequency (MAC)> 50. Individual protein levels (NPX) were inverse-rank normalized before analysis[8]. For the discovery cohort (n= 34,557), the model was adjusted for the following covariates: age, age², sex, age × sex, age² × sex, batch, UKB centre, UKB genetic array, time between blood sampling and measurement, and the first 20 genetic principal components[8].

The summary statistics of this data are available after applying for access through the UKB-PPP and adhering to the specified restrictions on use (https://www.synapse.org/Synapse:syn51364943/wiki/622119)[9].

***1.3 deCODE***

***1.3.1 Study participants***

Plasma samples were collected from 40,004 Icelanders between 2000 and 2019 through two primary initiatives: the Icelandic Cancer Project[10], which accounted for 52% of participants, and various genetic research programs at deCODE genetics in Reykjavík, Iceland, which contributed the remaining 48%[11]. For the Iceland Cancer Project, all Icelanders with prevalent and newly diagnosed cancer and their relatives were invited to participate in this study, together with a control population, randomly selected from the National Registry[11]. Though this sample set is enriched for cancer (36% correspond to a cancer diagnosis), the active cancer out of the whole set was below 1%[11]. For the samples collected at deCODE genetics, primarily came from the population-based deCODE Health study, with the remainder gathered through various other programs at deCODE[11]. Overall, the average participant age was 55 years (SD= 17 years), with 57% being women. Further details of the deCODE project are provided in the study by Ferkingstad et al.[11]

***1.3.2 Plasma Sample processing and quality control***

The plasma samples were analysed using the SomaScan version 4 assay (SomaLogic), which includes 5,284 aptamers that measure the relative binding of each plasma sample to the aptamers in relative fluorescence units[11]. In brief, the aptamers were transformed to recognize specific proteins, following the conversion of protein concentrations into corresponding DNA aptamer concentrations, and the levels of these proteins were quantified using a DNA microarray[11]. After quality control, 39,155 individuals of which 35,559 Icelanders were used in the protein GWASs[11]. Comparisons between Olink and SomaScan measurements were made in this study and a median correlation of 0.76 was found using 87 overlapping proteins in 199 Icelanders[11].

***1.3.3 Genotyping and imputation***

Whole genome sequencing (WGS) was performed in 49,708 Icelanders using Illumina technology[12]. A total of 166,281 Icelanders were genotyped with Illumina single-nucleotide polymorphism (SNP) chips, long-range phased and imputed based on the sequenced dataset, of which 35,559 had proteins measured by SomaScan version 4. Genotyping and imputation methods have been described in detail previously[13]. In this study, the analysis was restricted to variants with minor allele frequency (MAF) >0.01% and imputation information >0.9, resulting in 27.2 million imputed variants used in the GWAS[11].

***1.3.4 GWAS analyses***

For each of the 4,907 tested aptamers, rank-inverse normal transformed levels were adjusted for age, sex, and sample age separately for the deCODE Health study and the remaining studies[11]. The residuals were standardized using rank-inverse normal transformation, and the resulting standardized values were used as phenotypes for GWAS, utilizing the linear mixed model implemented in BOLT-LMM[11,14].

The summary statistics for this data are available upon request through deCODE Genetics and can be accessed for scientific purposes only (https://www.decode.com/summarydata/)[15].

***1.4 Fenland***

***1.4.1 Study participants***

The Fenland Study is a UK population cohort consisting of 12,435 participants born between 1950 and 1975, who were recruited from primary care in Cambridgeshire[16]. The recruitment of relatively young individuals, prior to the onset of chronic disease, was designed to investigate early processes and pathways leading to metabolic disease, unaffected by therapies or co-existing conditions[16]. Further details of the deCODE project are provided in the study by Pietzner et al.[17]

***1.4.2 Plasma sample processing***

Fasted EDTA plasma samples from 12,084 Fenland Study participants were collected at baseline and proteomic profiling was done by using by SomaLogic Inc. (Boulder, CO, USA) using an aptamer-based technology (SomaScan proteomic assay)[18]. The plasma abundances of 4,979 single-stranded oligonucleotides (aptamers), each with specific binding affinities to 4,775 distinct protein targets were measured (SomaLogic V4)[17]. After passing SomaLogic's quality control, only human protein targets were retained for further analysis (4,979 out of 5,284 aptamers)[18].

***1.4.3 Quality control, genotyping and imputation***

Three genotyping arrays were used for genotyping the Fenland participants, which includes: the Affymetrix UK Biobank Axiom array (OMICs, *N*=  8994), Illumina Infinium Core Exome 24 v1 (Core-Exome, *N*=  1060), and Affymetrix SNP 5.0 (GWAS, *N* =  1402)[18]. Variants with an MAF< 0.001, INFO< 0.4, or a Hardy-Weinberg equilibrium p-value < 10⁻⁷ in any of the genotyping subsets were excluded from further analyses[18]. Details of the QC, genotyping, and imputation procedures are provided in the study from Pietzner et al.[17]

***1.4.4 GWAS analysis***

After excluding ancestry outliers and related individuals, a protein-level GWAS was conducted on 10,708 participants of European descent from the Fenland Study[18]. A rank-based inverse normal transformation was applied to aptamer abundances to normalize their distribution[18]. The transformed abundances were then adjusted for age, sex, sample collection site, and 10 principal components. GWAS was performed using an additive model in BGENIE (v1.3)[18,19]. The results from the three genotyping arrays were combined in a fixed-effects meta-analysis using METAL[18,20]. Variants in the GWAS were restricted to those with a minor allele frequency of at least 1%. A total amount of 10,674 variant protein associations were identified.

The summary stats of this are available after applying for access with restricted use purposes (https://www.synapse.org/Synapse:syn51761394/wiki/622766)[21].

***1.5 OA GWA-meta data***

The largest genome-wide association meta-analysis (GWA-meta) of OA at present was conducted by Hatzikotoulas et al. and the summary results are available through the Musculoskeletal Knowledge Portal (https://msk.hugeamp.org/downloads.html)[22]. This GWA-meta study includes GWAS summary statistics from 87 GWAS summary statistics in 11 OA phenotypes, for a total of 1,962,069 individuals (ALLOA; Cases: 489,975; Controls: 1,472,094)[23]. In this study, 10 stratified OA phenotypes were defined, including hip and/or knee OA, knee OA, hip OA, total joint replacement, total knee replacement, total hip replacement, hand OA, finger OA, thumb OA, and spine OA. The detailed case–control counts for each phenotype are provided in Supplementary Table S14. OA was identified using various criteria, depending on the available data in these cohorts, such as self-reported OA, clinically diagnosed OA, ICD-10 codes, or radiographic evidence or joint replacement records. Controls were either OA-free or population-based, with or without ICD code or self-report exclusions.

***1.5.1 Quality control and meta-analysis***

The QC was performed centrally by using in-house scripts and EasyQC[20]. Missing data, mono-allelic SNVs, nonsensical values (p > 1, infinite beta’s etc.) and duplicates were removed from the data. Imputation were done for most of the GWAS using HRC[24] or 1000G[25] reference panel. The rest GWAS datasets (6 cohorts) were imputed by TOPmed or their own reference panel. All the data were harmonized with a standardized variant identifier across all datasets provided. Variants meeting the following criteria were excluded through using EasyQC: poor imputation quality (R^2^ < 0.3), if the effective sample size was < 20 and if the minor allele count was < 6. Allele frequency was checked by using HRC[24] or 1000G[25] reference panel as well. P-values were checked to correspond with their respective beta values, and the cleaned data were then used for meta-analysis. The meta-analysis was conducted using fixed-effect inverse variance weighting in METAL[20]. Genomic control was applied to all datasets, except those that had already been done[23]. A genome-wide significance threshold of p< 1.3×10⁻⁸ was set, correcting for multiple testing.

**2.1Analysis**

***2.1.1 MR analysis***

The outcome GWAS data were built on the hg19 genome version, and the analyses using UK Biobank Pharma Proteomics Project (UKBPPP) and Fenland data were also conducted based on the hg19 version. Since the GWAS data from deCODE are only based on hg38, the OA GWAS data were converted to the hg38 version using the LiftOver tool from the University of California Santa Cruz (UCSC) to align with the gene version[26]. Due to the presence of extreme beta values on the X chromosome in the OA outcome GWAS data, we restricted our analysis to autosomes only.

***2.1.2 Cis-MR analysis***

The cis-MR analysis was performed in parallel across three protein-wide GWAS databases as exposures, using data source from the Genetics of Osteoarthritis (GO) Consortium 2.0 as the outcome.

1. The human Major Histocompatibility Complex (MHC) region (chr6: from 26Mb to 34Mb) was firstly removed for each protein from 3 databases due to the complex Linkage disequilibrium (LD) structure of SNPs within[27].
2. Only the cis-region (± 200kb of each gene position) of protein quantitative trait loci (pQTLs) was selected for valid SNP identification.
3. SNP(s) that were significantly (*p*< 5 × 10^-6^) associated with all proteins were then selected.
4. A QC step was applied to exclude instrumental variables (IVs) with MAF < 0.1%.
5. Single- nucleotide variant (SNVs) identified by the previous step were selected from each type of OA.
6. LD clumping for those selected SNV(s) was performed using the European super-population from the 1000 Genomes phase 3 reference panel, applying the PLINK[28] to identify independent pQTL(s) for each protein. *r*^2^< 0.1 and clump kb=10 kb was used to exclude the dependent pQTL(s). Due to precision limits in floating-point representation that resulted in some identical p-values among IVs, pseudo p-values were generated for PLINK clumping. These were calculated by deriving absolute p-values from the beta and standard error values, using the z-score and computing the –log₁₀(p) via the pnorm() and mpfr() functions from the Rmpfr package (version 1.0-0).
7. Prior to conducting the MR analysis, the LD matrix of r values (correlation coefficients) was calculated using the European super-population from the 1000 Genomes phase 3 reference panel and the ‘ieugwasr’ R package (version 1.0.2)[29].
8. For single IVs, MR Wald ratio methods was applied by using ‘TwoSampleMR’ R package (version 0.6.9)[30,31]. For a number of IVs over 2, Inverse-variance weighted analysis was performed by using ‘MendelianRandomization’ R package (version 0.10.0)[32] and the correlation (LD) matrix was considered in the analysis.
9. Benjamini-Hochberg (B-H) procedure[33] were used for multiple test correction for MR analysis.

***2.1.3 Colocalisation analysis***

Results that passed the B-H procedure were again chosen to perform a Bayesian colocalisation analysis[34]. This analysis aimed to estimate the posterior probability that each genomic locus contains a single variant affecting both the protein and each OA trait, rather than the variant being shared coincidentally due to LD. The ‘coloc’ R package (version 5.2.3) was used in this section[34,35]. There are 4 hypotheses in colocalisation analysis which include: *H_0_*: neither trait has a genetic association in the region; *H_1_*: only trait 1 has a genetic association in the region; *H_2_*: only trait 2 has a genetic association in the region; *H_3_*: both traits are associated, but with different causal variants; *H_4_*: both traits are associated and share a single causal variant. Among the four hypotheses, protein-OA pairs with PPH_4_ > 80% were deemed to have colocalisation evidence in this study.

***2.1.4 Reverse MR***

To assessed the possibility of reverse causation, results that passed the B-H procedure[33] from the cis-MR analysis were used to perform a reverse MR analysis:

1. The MHC region for each protein across three databases was also removed.
2. SNV(s) that was/were significantly (P-value ≤ 5 × 10^-6^) associated with each OA type were selected.
3. SNV(s) identified by the previous step were selected for each protein from 3 databases.
4. The Steiger filtering analysis[30] was performed among all the IVs before the MR analysis.
5. The same MR analysis was performed as described in the ‘Cis-MR analysis’ section.
6. B-H procedure was performed among MR results.

***2.1.5 Quality control and Differential protein expression analysis***

We conducted a differential protein expression (DPE) analysis using individual-level proteomics data from a UK Biobank sub-cohort comprising participants with prevalent OA (cases) and those without OA before the assessment date (control), who attended the initial assessment visit between 2006 and 2010, across 11 OA phenotypes. QC was performed, accounting for the missingness of the OLINK protein biomarkers dataset. Only data from participants at baseline visits were selected. Proteins with equal and over 30% missingness value and participants with equal and over 30% missingness value were removed from the datasets. The number of cases and prevalent OA cases after QC are provided in Supplementary Table S15. Imputation of covariates are implemented on body mass index (BMI), Townsend deprivation index (TDI) and smoking status. BMI obtained from by impedance measurement[36] was used and the missing values were first replaced by using BMI value that constructed from height and weight measured[37]. The rest of the BMI missing values, smoking status, and TDI at recruitment were imputed using age, gender, and the top 10 principal components of genetic ancestry as predictors in the multiple imputation (MI) process with the MICE R package (version 3.17.0)[38]. Continuous variable and categorical variable were imputed using predictive mean matching (pmm) and polynomial regression (polyreg) models respectively. The protein data was imputed by using impute.knn() function in the impute package (version 1.80.0) in R[39]. DPE analysis was done for each protein and OA phenotype. In the primary analysis, each protein was iteratively regressed using age, sex, and imputed BMI as covariates. The B-H procedure[33] ($P_{adjust}=\frac{P_{current}\times m}{i}$ ; *P*_current_ is the original p-value; m is the total number of hypotheses; *i* is the rank of the current *P*-value.) was applied to control the false discovery rate (FDR). Sensitivity analyses were performed by including the top 3 principal components of genetic ancestry, smoking status, and TDI as additional covariates in the main analysis.

In addition to the methods described in the main text, we converted the coefficient of variation (CV) formula provided in the supplementary materials of Sun et al.[6] into an minimum detectable difference (MDD) formula. Using the minimum intra-person CV (1.8%) observed in Olink proteins, we calculated the lowest detectable MDD threshold as 0.051 and applied it to filter the beta estimates, excluding regression results with beta ≤ 0.051. The MDD was calculated using the following function:

$${MDD}_{NPX}=2^{z\cdot\sqrt{2}\cdot\frac{1}{ln2}\cdot\sqrt{ln(1+{(\frac{CV}{100})}^{2})}}-1$$

**3.1 Biological insights**

***3.1.1 PPI network***

The STRING (Search Tool for the Retrieval of Interacting Genes) database (https://string-db.org/)[40] provides comprehensive information on both physical protein interactions and functional protein associations. The data within STRING are sourced from a variety of methods, including automated text mining of scientific literature, computational predictions based on co-expression and conserved genomic context, experimental interaction databases, and curated pathways or complex datasets. A group of genes encoding the proteins analysed from MR significant proteins, MR significant proteins from UKBPPP, and significant proteins from DPE analysis were used to construct protein-protein interaction (PPI) networks to investigate how one protein interacts with others intracellularly by using the STRING, v11.5 [41]. PPI results were obtained from STRING, and the CytoNCA plugin in Cytoscape was used to calculate eight standard centrality metrics based on the PPI network: Betweenness Centrality (BC), Closeness Centrality (CC), Degree Centrality (DC), Eigenvector Centrality (EC), Local Average Connectivity-based method (LAC), Network Centrality (NC), Subgraph Centrality (SC), Information Centrality (IC)[42]. Specifically, BC measures how often a node lies on the shortest paths between other nodes; CC measures the average shortest path length from a node to all others; DC quantifies the number of direct neighbours of a node; EC assigns greater importance to nodes connected to other highly connected nodes (via the eigenvectors of the adjacency matrix); LAC evaluates how well a node’s neighbours are connected among themselves; NC accounts for both a node’s neighbours and their common neighbours; SC quantifies the extent to which a node participates in closed walks (subgraphs) of all sizes using adjacency matrix powers; and IC assesses the reduction in overall network efficiency when a node is removed[42]. Median selection was applied to the PPI results, and a subnetwork was constructed in STRING using proteins that exceeded the median value across all eight centrality measures.

***3.1.2 Over-representation analysis***

Three over-representation analyses were conducted by using a group set of genes derived from MR significant proteins, MR significant proteins from UKBPPP, and significant proteins from DPE analysis using the EnrichR (https://maayanlab.cloud/Enrichr/)[43]. To be specific, we used the Gene Ontology (GO) knowledgebase (http://geneontology.org), which serves as a comprehensive resource for understanding the functions of genes and their products[44–46]. The GO is based on the "molecular biology paradigm," which categorizes gene function into three main aspects: Molecular Function (MF), Cellular Component (CC), and Biological Process (BP)[45]. For pathway analysis, we used the WikiPathways 2024 database (https://www.wikipathways.org/), which reportedly contains a significantly larger number of pathways than the combined collections of WikiPathways, Reactome, and KEGG[47]. The Drug Signature Database (DsigDB) was used to identify approved drugs and compounds that interact with the identified proteins and to predict drug candidates for OA treatment[48].

***3.1.3 Druggability assessment***

Druggability profiles for proteins with stringent colocalisation evidence were obtained from a previously published list of druggable genes[49]. The gground[50] and ggprism[51] r packages were used for data visualisation.
