## Supplementary Fig. for "Genetically informed search for potential osteoarthritis drug targets across the proteome"

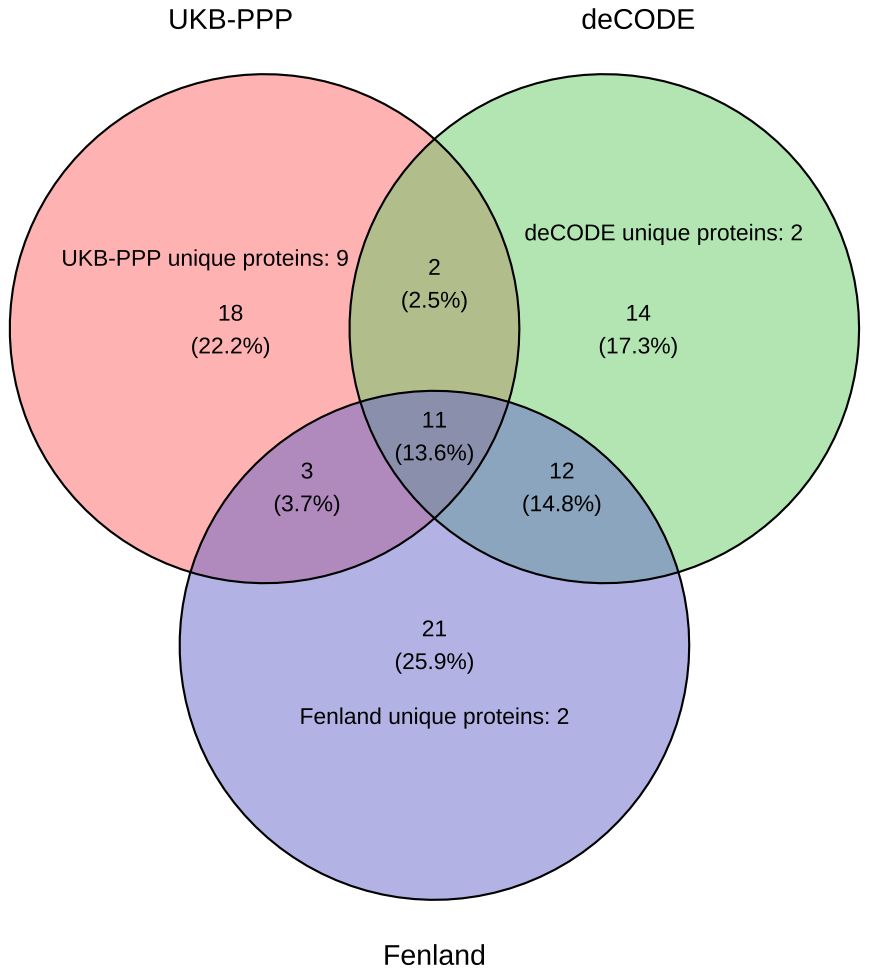

**Supplementary Fig. 1.** Venn diagram illustrating the overlap of MR results across the three cohorts. Numbers indicate proteins unique to each cohort or shared between cohorts. MR, mendelian randomisation; UKB-PPP, UK Biobank Pharma Proteomics Project.

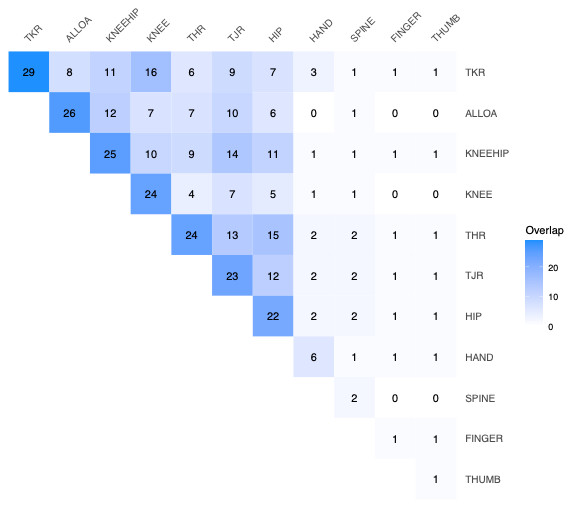

**Supplementary Fig. 2.** Heatmap of pairwise overlaps in MR significant proteins across 11 OA phenotypes. The numbers within the cells indicate the number of proteins shared between each pair of OA types, with color intensity reflecting the degree of overlap. MR, mendelian randomisation; OA, osteoarthritis; TJR, total joint replacement; THR, total hip replacement; TKR, total knee replacement.

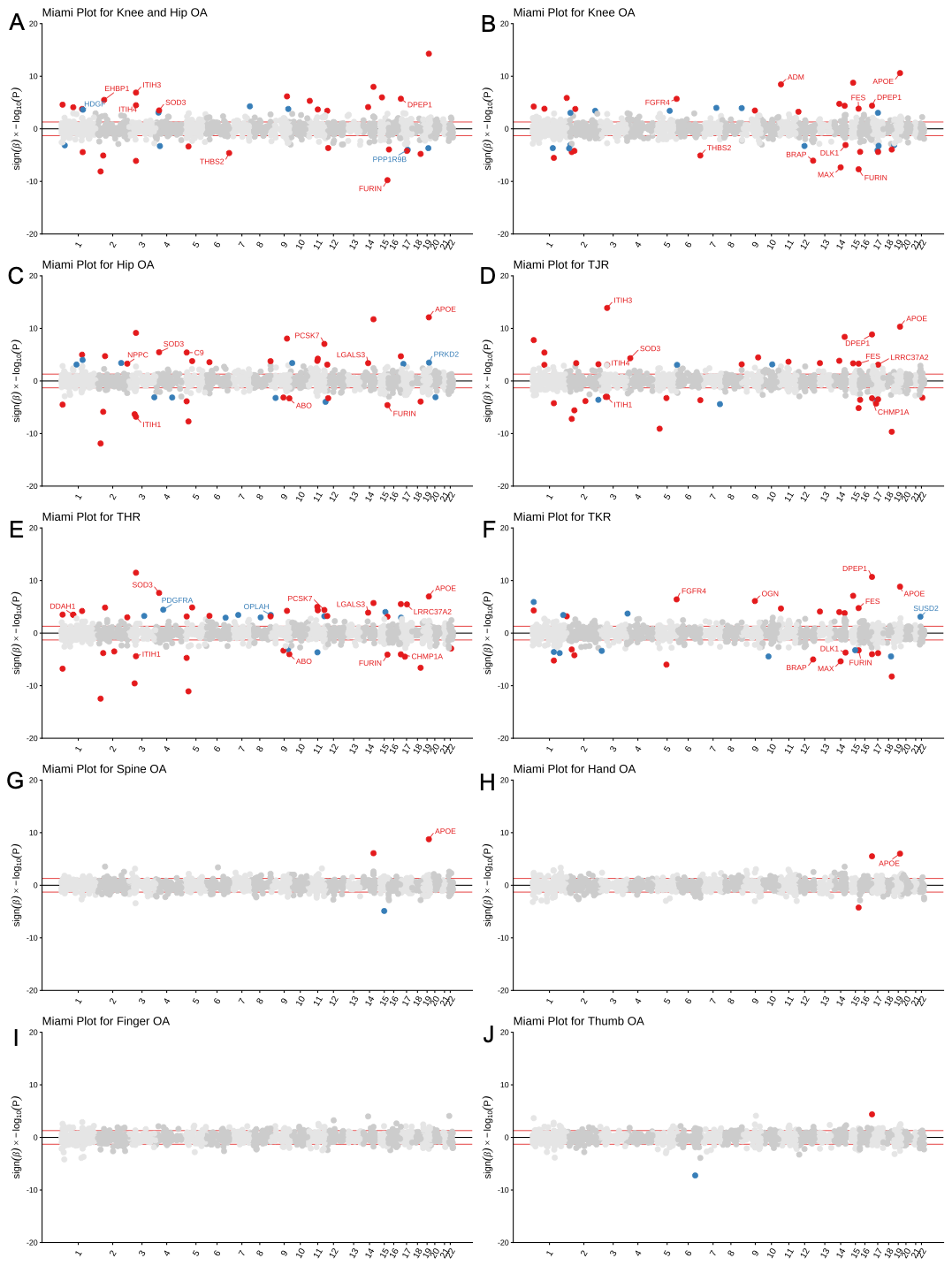

**Supplementary Fig. S3.** Miami plots of MR results across 10 OA (Supplementary Fig. S3A-S3J) phenotype in the UKB-PPP data source. The x-axis shows the chromosomal position of each protein’s encoding gene, and the y-axis represents the signed effect estimate (β) multiplied by the negative log10-transformed p-value after Benjamini–Hochberg correction, allowing visualization of effect direction. The red horizontal lines indicate the nominal significance threshold (p = 0.05). Proteins highlighted in blue indicate MR associations specific to the current OA phenotype in current data sources, while those in red are supported by MR evidence in two or more phenotype. Proteins fulfilling colocalisation criteria (PPH_4_ > 0.8) are labeled. MR, mendelian randomisation; UKB-PPP, UK Biobank Pharma Proteomics Project; OA, osteoarthritis; PPH_4_, posterior probability of hypothesis 4; TJR, total joint replacement; THR, total hip replacement; TKR, total knee replacement.

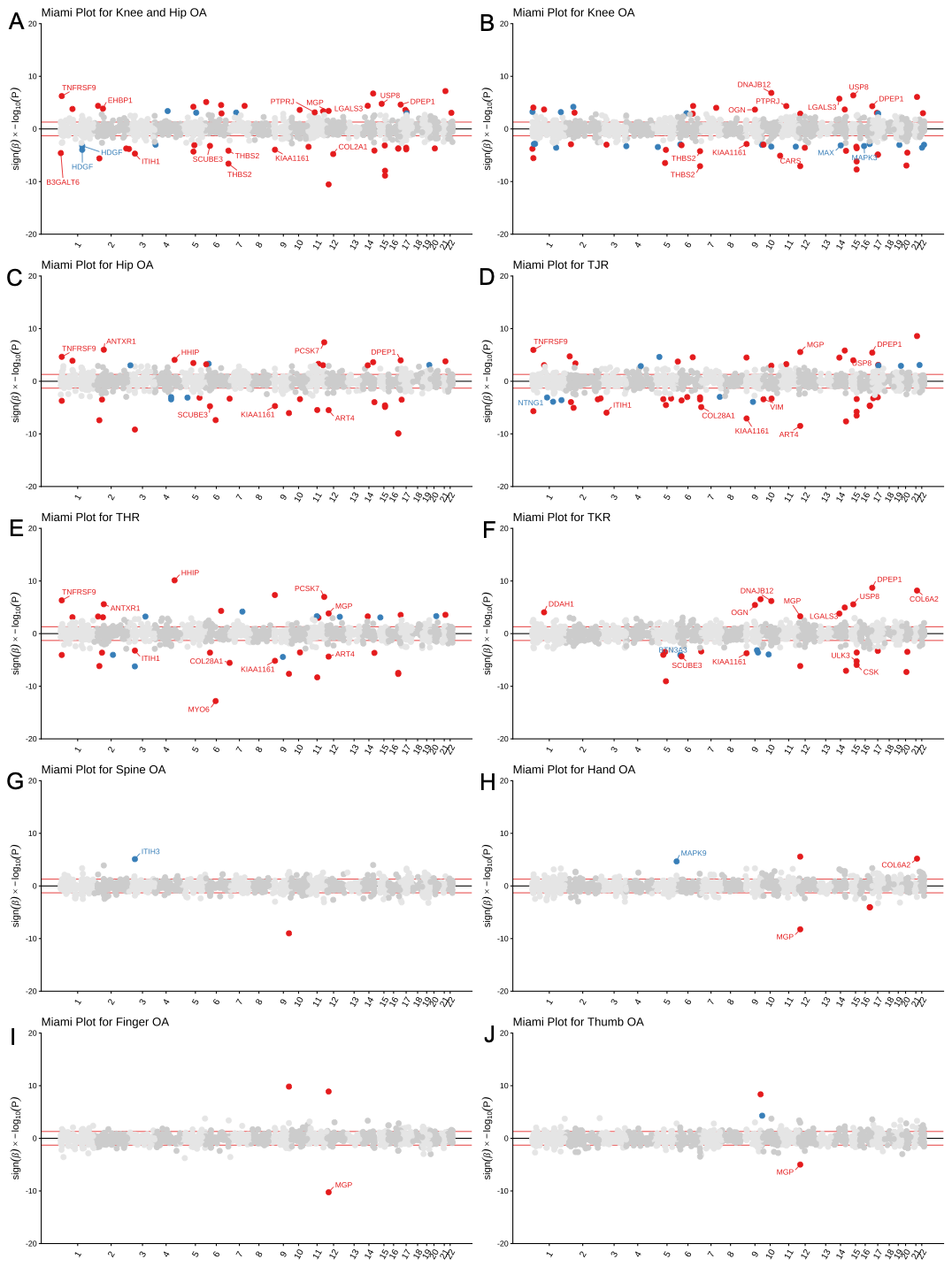

**Supplementary Fig. S4.** Miami plots of MR results across 10 OA phenotypes (Supplementary Fig. S4A-S4J) in the deCODE data source. The x-axis shows the chromosomal position of each protein’s encoding gene, and the y-axis represents the signed effect estimate (β) multiplied by the negative log10-transformed p-value after Benjamini–Hochberg correction, allowing visualization of effect direction. The red horizontal lines indicate the nominal significance threshold (p = 0.05). Proteins highlighted in blue indicate MR associations specific to the current OA phenotype in current data sources, while those in red are supported by MR evidence in two or more phenotype. Proteins fulfilling colocalisation criteria (PPH_4_ > 0.8) are labeled. MR, mendelian randomisation; UKB-PPP, UK Biobank Pharma Proteomics Project; OA, osteoarthritis; PPH_4_, posterior probability of hypothesis 4; TJR, total joint replacement; THR, total hip replacement; TKR, total knee replacement.

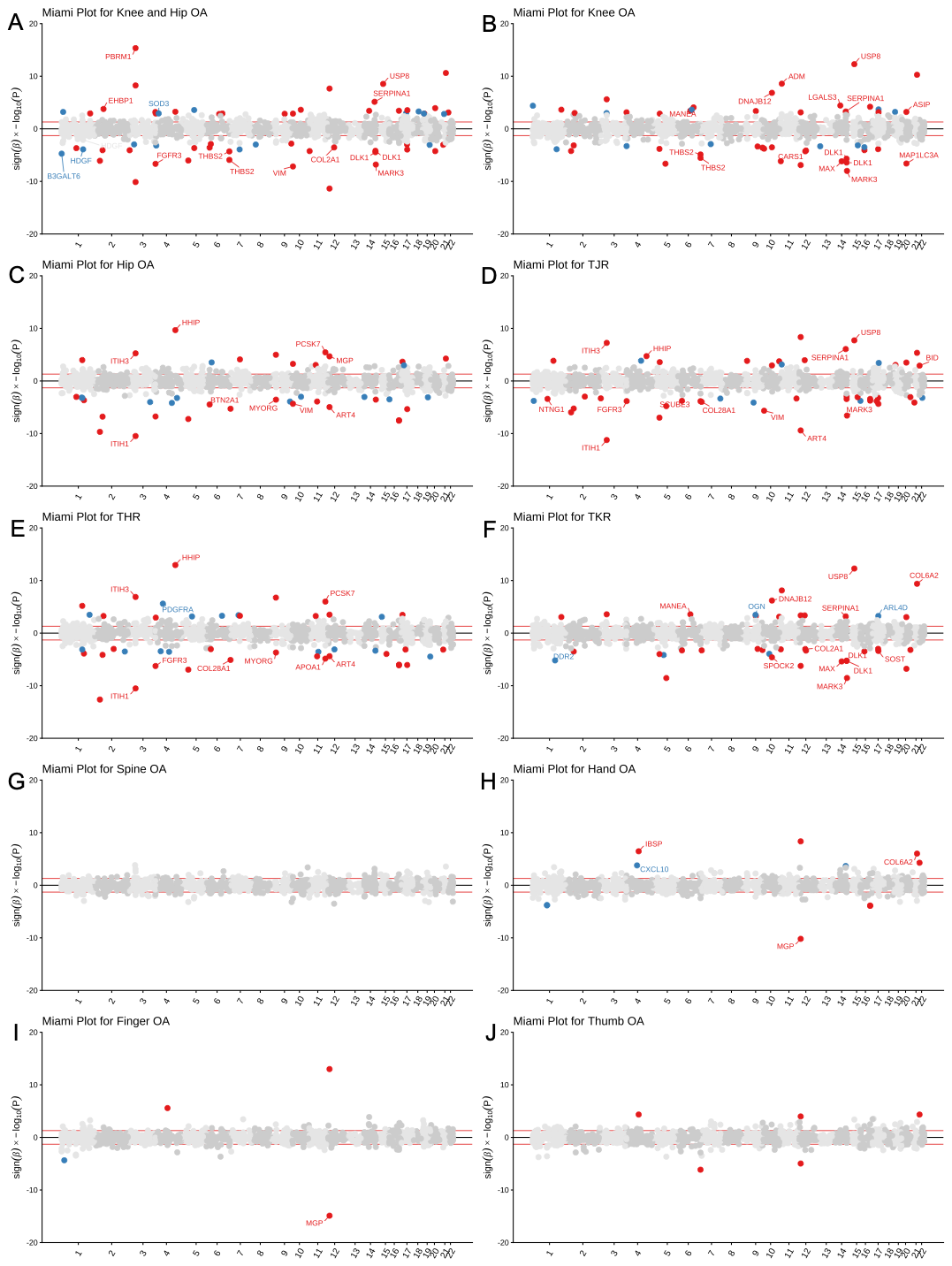

**Supplementary Fig. S5.** Miami plots of MR results across 10 OA phenotypes (Supplementary Fig. S5A-S5J) in the Fenland data source. The x-axis shows the chromosomal position of each protein’s encoding gene, and the y-axis represents the signed effect estimate (β) multiplied by the negative log10-transformed p-value after Benjamini–Hochberg correction, allowing visualization of effect direction. The red horizontal lines indicate the nominal significance threshold (p = 0.05). Proteins highlighted in blue indicate MR associations specific to the current OA phenotype in current data sources, while those in red are supported by MR evidence in two or more phenotype. Proteins fulfilling colocalisation criteria (PPH_4_ > 0.8) are labeled. MR, mendelian randomisation; UKB-PPP, UK Biobank Pharma Proteomics Project; OA, osteoarthritis; PPH_4_, posterior probability of hypothesis 4; TJR, total joint replacement; THR, total hip replacement; TKR, total knee replacement.

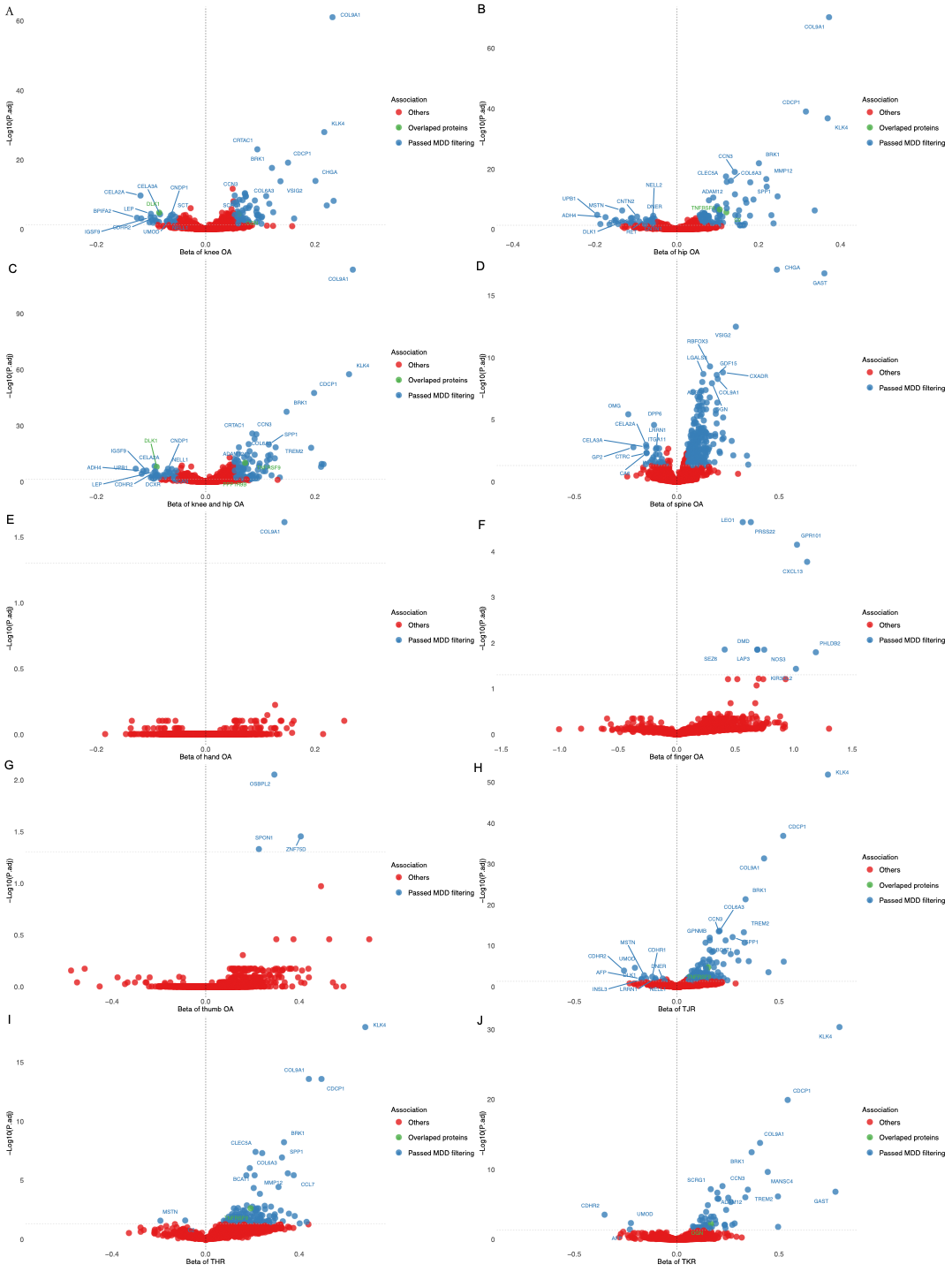

**Supplementary Fig. S6.** Volcano plots of DPE results across 10 OA phenotypes (Supplementary Fig. S6A-S6J). The x-axes show beta coefficients representing the effect size of each protein on risk of the indicated OA phenotype, and the y-axes show –log10 of the adjusted p-values from linear regression. Proteins passing the MDD threshold (Beta > 0.051) are shown in blue. Proteins overlapping between DPE and MR analyses across three proteomic datasets with matched OA types are shown in green. The top 10 most significant proteins on each side of the beta distribution are labelled. Other proteins are displayed in red. DPE, differential protein expression; OA, osteoarthritis; MDD, minimum detectable difference; MR, mendelian randomisation; TJR, total joint replacement; THR, total hip replacement; TKR, total knee replacement.

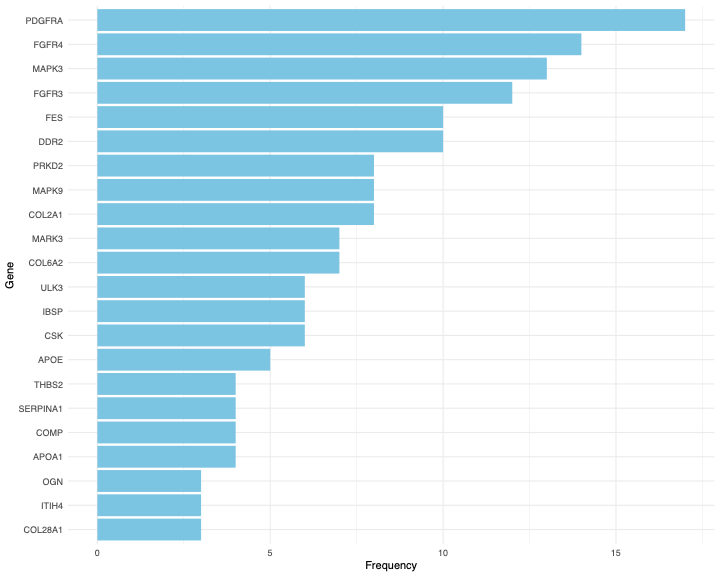

**Supplementary Fig. S7.** Frequency plot of the top 20 genes identified across the top five over-representation analysis results from MR-significant proteins. The x-axis shows the frequency of each gene, and the y-axis lists the corresponding genes. MR, mendelian randomisation.

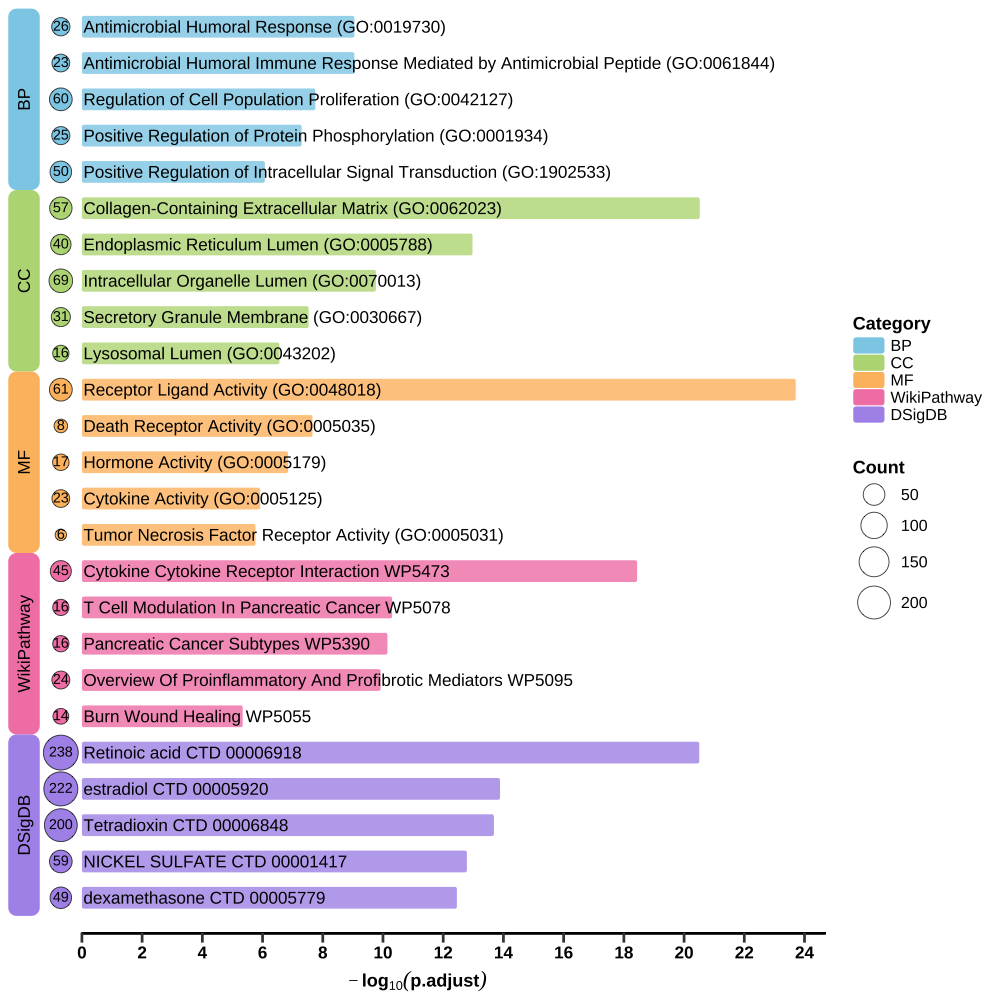

**Supplementary Fig. S8.** Top five over-representation analysis results based on 605 significant proteins in DPE analysis, using GO, WikiPathway, and DsigDB databases. Categories include BP, CC, MF, and DsigDB. Circle sizes represent the number of proteins enriched in each pathway or term. DPE, deferential protein expression; GO, Gene Ontology; DsigDB, Drug Signature Database; BP, biological process; CC, cellular component; MF, molecular function.

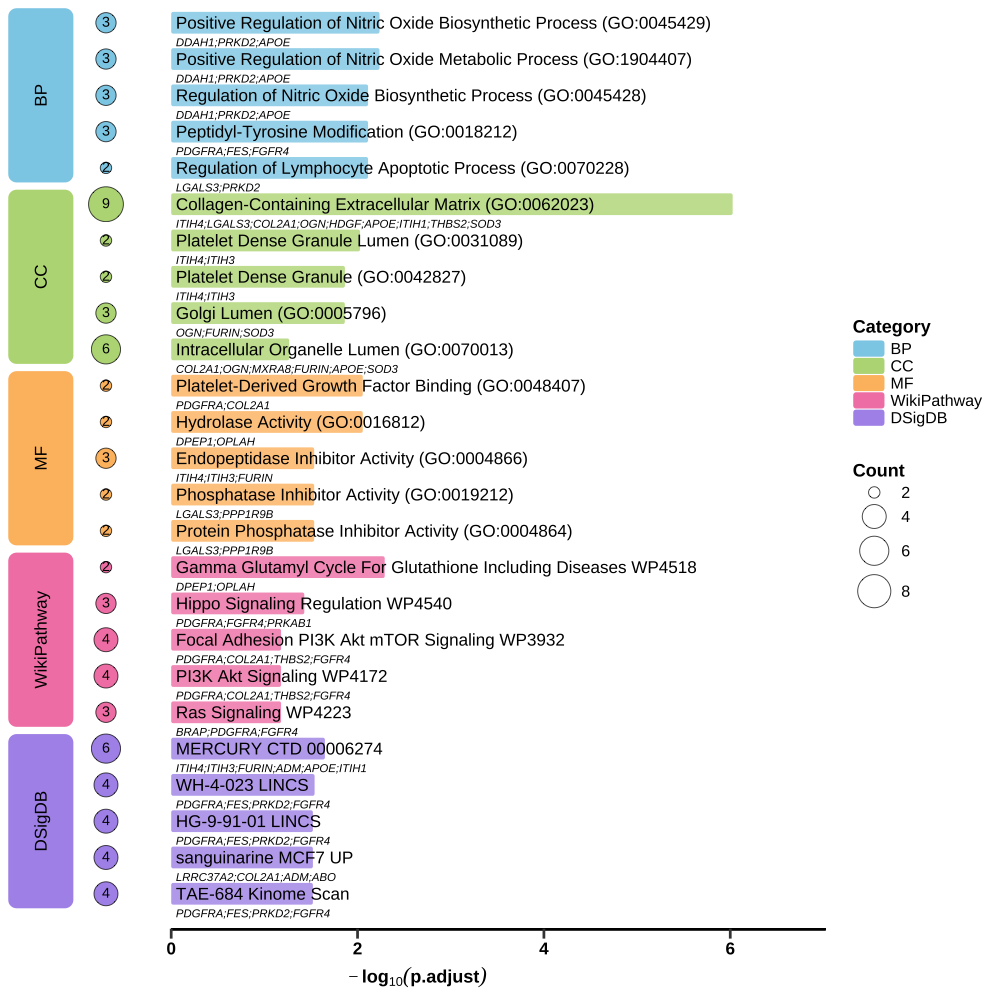

**Supplementary Fig. S9.** Top five over-representation analysis results based on 34 MR significant proteins from UKB-PPP, using GO, WikiPathway, and DsigDB databases. Categories include BP, CC, MF, and DsigDB. Circle sizes represent the number of proteins enriched in each pathway or term. MDD, minimum detectable difference; DPE, deferential protein expression; GO, Gene Ontology; BP, biological process; CC, cellular component; MF, molecular function; DsigDB, Drug Signature Database.

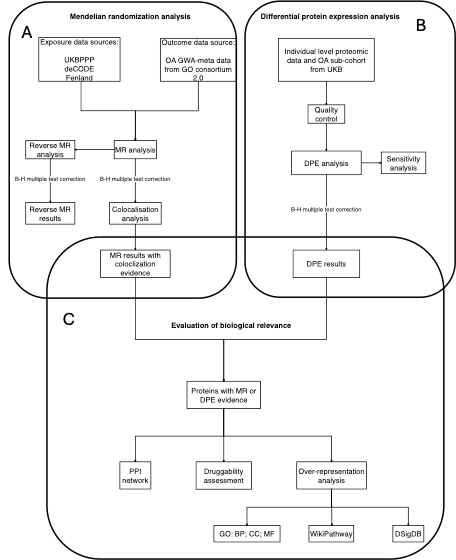

**Supplementary Fig.** **S10.** Flowchart summarizing the overall study design. MR, mendelian randomisation; DPE, differential protein association; OA, osteoarthritis; UKB, UK Biobank; B-H, Benjamini–Hochberg; UKB-PPP, UK Biobank Pharma Proteomics Project; meta-GWAS, meta-analysis of genome-wide association studies; GO consortium 2.0, Genetics of Osteoarthritis Consortium 2.0; GO, Gene Ontology; BP, biological process; CC, cellular component; MF, molecular function; DSigDB, Drug Signature Database.
